# Is exercising better for your Mental Health?

**DOI:** 10.1101/2023.03.28.23287187

**Authors:** Dev Himanshubhai Desai, Aparajita Shukla, Himanshu Desai

## Abstract

**Objective:** To correlate psychological changes with exercise.

**Methodology:** This cross sectional study was conducted on 102 participants in a known Gym with age group 17-75 (28.3 ± 10.4, 70.59% males). They were given **“Rosenberg self-esteem Questionnaire”, “Satisfaction with life scale” & “Hamilton & Beck’s Depression Inventory” in** pen-paper type and were asked to fill it twice; once at the present time and once in retrospection before they started doing exercise. Three different values of Self-esteem, Satisfaction and Depression were collected. Answers from these Depression, Self-esteem questionnaire and Satisfaction questionnaire were summed up to get the final individual score.

**Results:** Overall gain was 62.53% suggesting positive impact of exercise. Highest impact was seen in age group of 17-30 of 72.46%. Lower education groups showed significantly higher gain, compared to higher educational groups (191.80%vs42.64%,p<0.05), which may be because of already higher scores in pre-exercise stage among higher education group. Target-achievers showed significantly higher gain compared to non-target-achievers (84.84%vs33.18%,p<0.01)).

**Conclusion:** Exercise has an overall positive impact on satisfaction, self-esteem and reduction in depression. People are influenced by different factors into getting fit and most of them agree that body image has a value in their perception of any individual.

## Introduction

It is pretty evident that perception of one’s self as their own body image has a substantial influence on their mentality and their psychology and affects their daily routine. (1) Normally weight issues and body image have always been associated to mental factors like depression, anxiety, stress etc. (2)

Mental Health in recent times is emerging as one of the most important if not the most important aspect of overall health. Good mental health helps individual achieve success which a person with similar caliber but poor mental health cannot achieve. (3)

There are substantial evidences pointing out the fact that mental health and physical health are not two separate entities but are correlated and associated heavily and both affect each other vastly, more than we believe. (4-7)Deprived mental health in turn leads to inactivity and therefore increased deterioration of physical health. (2) Inversely to that physical activity has been found individually be stimulating and as a potent Stress-Buster. (8)Physical activity has also been found to have lowering effect on levels of Depression and its helpful in increasing Self-esteem of a person by being satisfied with oneself and feeling of acceptance in one’s social circle. (9)

Achievement of desired target weight has shown to have releasing neurotransmitters which causes increased self-confidence and decreased depression somewhat better than just exercising and not achieving target in given time as it gives a feeling of accomplishment boosting the self-acceptance and satisfaction levels. (9)

There are many existing arguments about increasing quality of physical health to increase quality of mental health. (10) A sound mind and a sound body go hand in hand. (11) (12) And as it is said that the way of mental fitness has multiple paths including physical fitness, it is important to see how potent this path is. It needs to be assessed if the increase in mental health is as substantial as we think it to be and if yes, can we use it as a regular mode of therapy to combat gruesome mental health problems. (13) (14) (15)

## Methods and materials

A cross sectional Retrospective Pre-Post analysis was carried out. The data was collected at a local gym where people come to exercise and increase their overall physical health. A questionnaire was formed combining **“Rosenberg self-esteem Questionnaire”**(16) (17) (18), **“Satisfaction with life scale”** (19, 20) **& “Hamilton & Beck’s Depression Inventory”** (21)in pen-paper type and participants were asked to fill it twice; once at the present time and once in retrospection before they started doing exercise. Demographic details of those participants were also obtained to co-relate the scores from the scale with age, gender, education and whether they have achieved their target or not. Along with the scales and the demographic details the participants had questions about who influenced them and whether they think that body image is an important factor in their perception about any fellow human being. Data collection was done individually after explaining the purpose and the method of the study.

As the study was survey based, oral exemption from ethics committee was gained before the study. The Authors have no conflict of interests nor their received any funding or financial gain for the study.

### Inclusion criteria

- Participants who were willing to give consent to fill the form
- Participants who are regularly coming to the gym to exercise since last 6 months or more

### Exclusion criteria

- Participants who were not willing to fill the form
- Participants with existing medical condition
- Participants with mental disorder
- Participants with metabolic diseases

### Scales

#### “Rosenberg self-esteem Questionnaire”

This 10 item questionnaire is the most popular tool for measuring self-esteem. Rosenberg Self-Esteem Scale (RSES), (1965) is a one-dimensional tool which concentrates on the perception of self-esteem an individual has for their own value. The questionnaire consists of 10 questions, 5 positive and 5 negative graded statements. This Rosenberg self-esteem scale, developed by the sociologist Morris Rosenberg. (16) (17) (18)

Here it was used to estimate the change in self-esteem and thus changes in total score.

#### “Satisfaction with life scale”

This 5-item questionnaire is scored on 7 variable answers. The Satisfaction with Life Scale**(SWLS)** was developed to assess satisfaction with people’s lives as a whole. The scale allows subjects to integrate and weigh satisfaction with specific life domains, such as health or finances in whatever way they choose. The Satisfaction With Life Scale has been used heavily as a measure of the life satisfaction component of subjective well-being. Scores on the SWLS have been shown to correlate with measures of mental health, and be predictive of future behaviors such as suicide attempts. (19, 20)

Here it was used to see the satisfaction of participants with themselves and changes in the level of satisfaction.

#### “Hamilton & Beck’s Depression Inventory”

This questionnaire is widely used to measure levels of depression in patients. It is validated by a study conducted by Storch, Roberti and Roth (2004) (22) examined the psychometric properties of the BDI-II (21). 414 students from two separate universities participated in the study. Their results “supported BDI-II two-factor structure measuring cognitive-affective and somatic depressive symptoms”. It is preferably used to check depression over other questionnaire due to its high beneficence.

### Data Entry and analysis

The data was entered in Microsoft Excel. Qualitative analysis in the form of Averages, percentage gain, and other statistical methods like Paired T-test were used to establish correlations between scores and other variables included in the study questionnaire.

The analysis had three different values of Self-esteem, Satisfaction and Depression. As the depression was a negative questionnaire, it’s score were substituted from the summation of scores from Self-esteem questionnaire and Satisfaction questionnaire.

Hence,

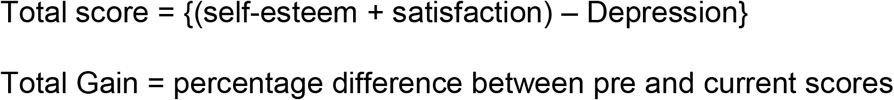

This total score was then correlated with Age, Gender, level of education and target achievers.

## Results

### Participants and Demography

A total of 102 participants took part in this cross sectional study in which 29.41%(30) were Females whereas 70.59%(72) were Male participants.

30.39%(31) were Married whereas 69.61%(71) were Unmarried participants.

Participants’ age group with their number and in percentage (n=102) (table 1)

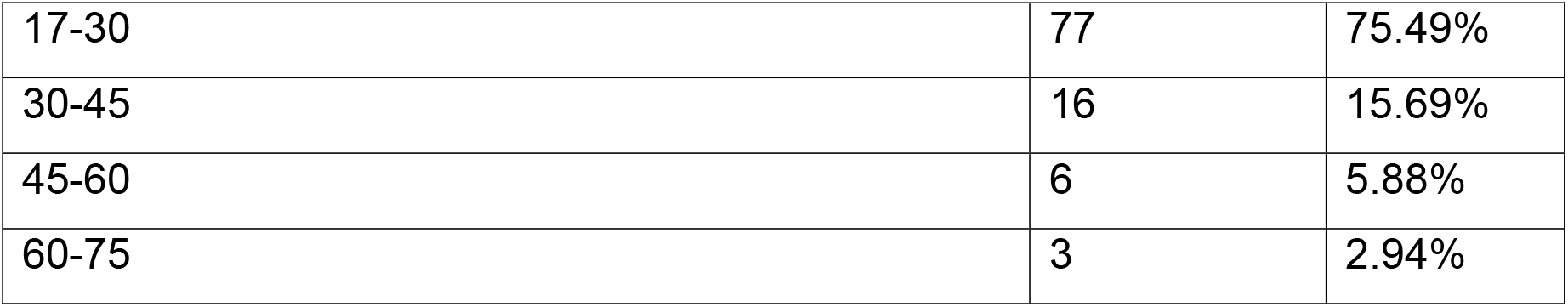

Participants’ level of education (n=102) (table 2)

Participants' level of education (n=102) (table 2)
|  |  |  |
| --- | --- | --- |
| PRIMARY | 0 | 0.00% |
| SECONDARY | 6 | 5.88% |
| HIGHER SECONDARY | 6 | 5.88% |
| GRADUATE | 63 | 61.76% |
| POST GRADUATE | 27 | 26.47% |

Overall changes in Total score (n=102) (table 3)

| Before | Present time | Difference % | P value |
| --- | --- | --- | --- |
| 30.48 ± 18.94 | 49.59 ± 13.69 | 62.53 | <0.0001 |

The Total score changes by Age (n=102) (table 4)

|  | Before | Present | Difference | N | P value |
| --- | --- | --- | --- | --- | --- |

|  |  | time | % |  |  |
| --- | --- | --- | --- | --- | --- |
| 17-30 | 28.10 ± 18.39 | 48.47 ± 13.26 | 72.46 | 77 | <0.0001 |
| 30-45 | 32.19 ± 20.21 | 51.13 ± 16.09 | 58.84 | 16 | =0.0064 |
| 45-60 | 43.33 ± 11.72 | 53.33 ± 13.35 | 23.08 | 6 | =0.198 |
| 60-75 | 56.67 ± 10.21 | 62.67 ± 5.50 | 10.59 | 3 | =0.421 |

The Total score changes in relation with Gender (n=102) (table 5)

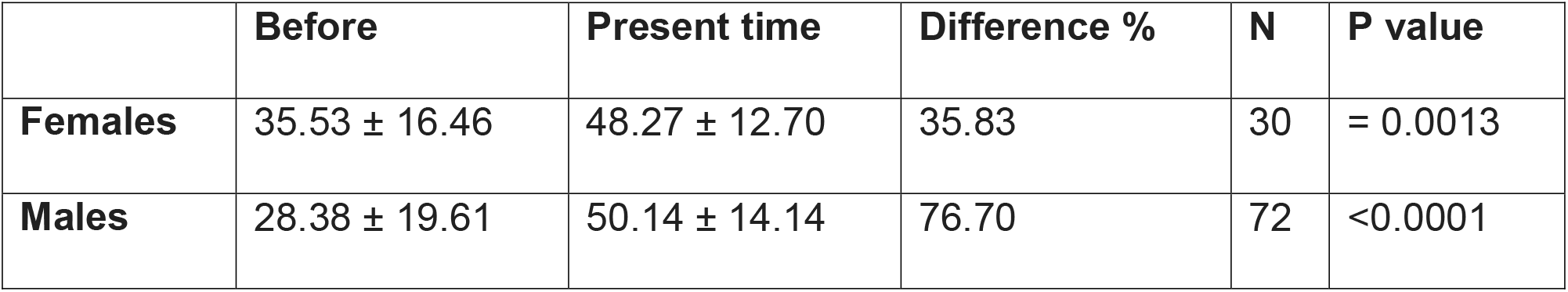

The Total score changes by level of Education (n=102) (table 6)

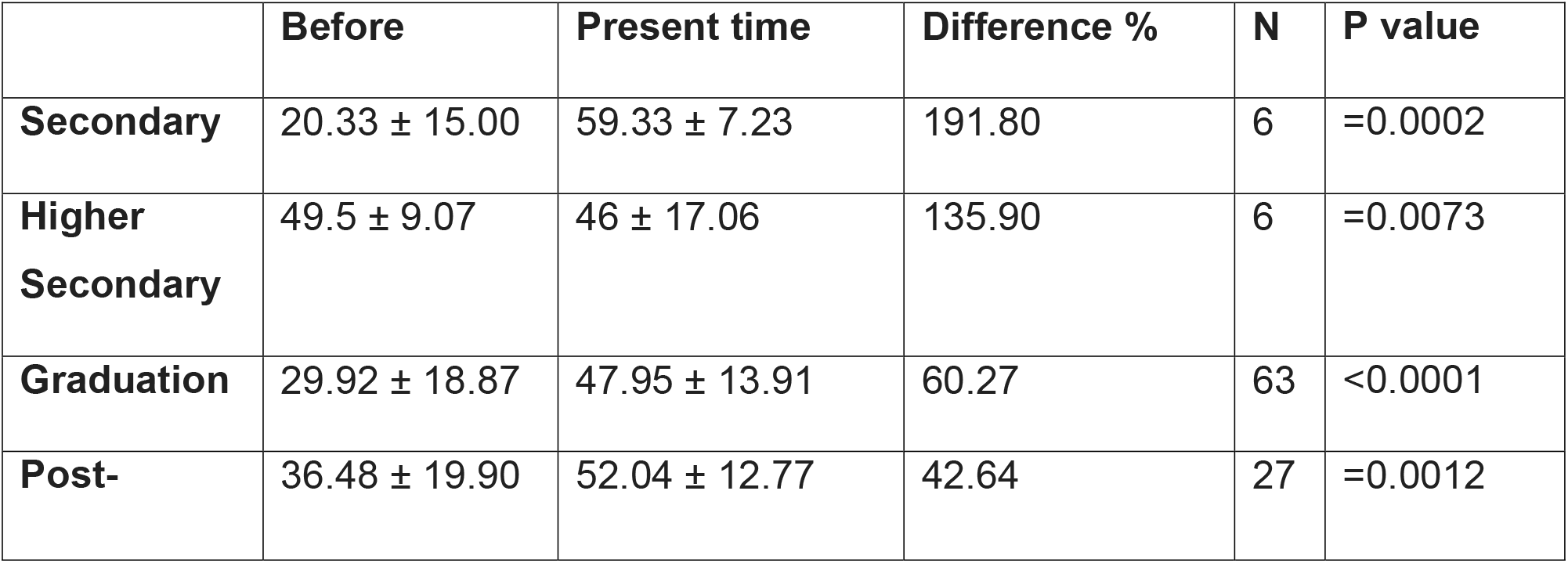
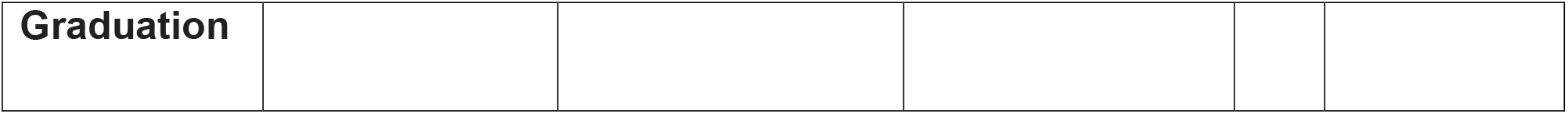

|  | **Before** | **Present time** | **Difference %** | **N** | **P value** |
| --- | --- | --- | --- | --- | --- |
| **Secondary** | 20.33 ± 15.00 | 59.33 ± 7.23 | 191.80 | 6 | =0.0002 |
| **Higher Secondary** | 49.5 ± 9.07 | 46 ± 17.06 | 135.90 | 6 | =0.0073 |
| **Graduation** | 29.92 ± 18.87 | 47.95 ± 13.91 | 60.27 | 63 | <0.0001 |
| **Post-** | 36.48 ± 19.90 | 52.04 ± 12.77 | 42.64 | 27 | =0.0012 |
| **Graduation** |  |  |  |  |  |

The Total score difference in just exercising and Target achievers (reduced weight by 10% / gained weight by 5% / are body-builders) (n=102) (table 7)

|  | **Before** | **Present time** | **Difference %** | **N** | **P value** |
| --- | --- | --- | --- | --- | --- |
| **Target Achievers** | 26.92 ± 19.06 | 49.76 ± 13.78 | 84.84 | 66 | <0.0001 |
| **Others** | 37 ± 17.13 | 49.28 ± 13.55 | 33.18 | 36 | =0.0012 |

Influencers who influenced participants (chart 1)

(a question was asked to participants about who influenced them most in their life)

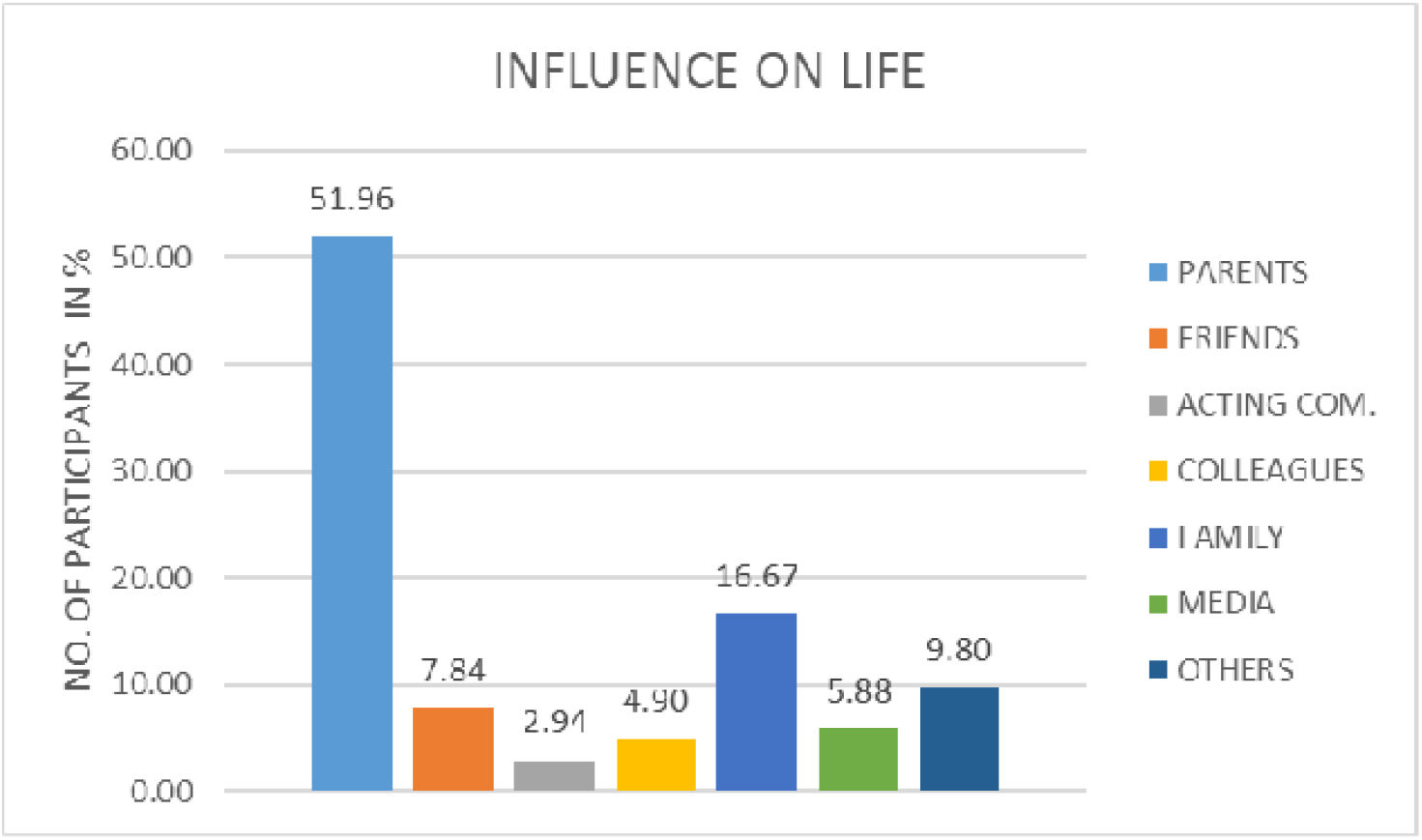

Body image (chart 2)

(a question was asked to participants if they thought that body shape is an important factor in their perception of anybody)

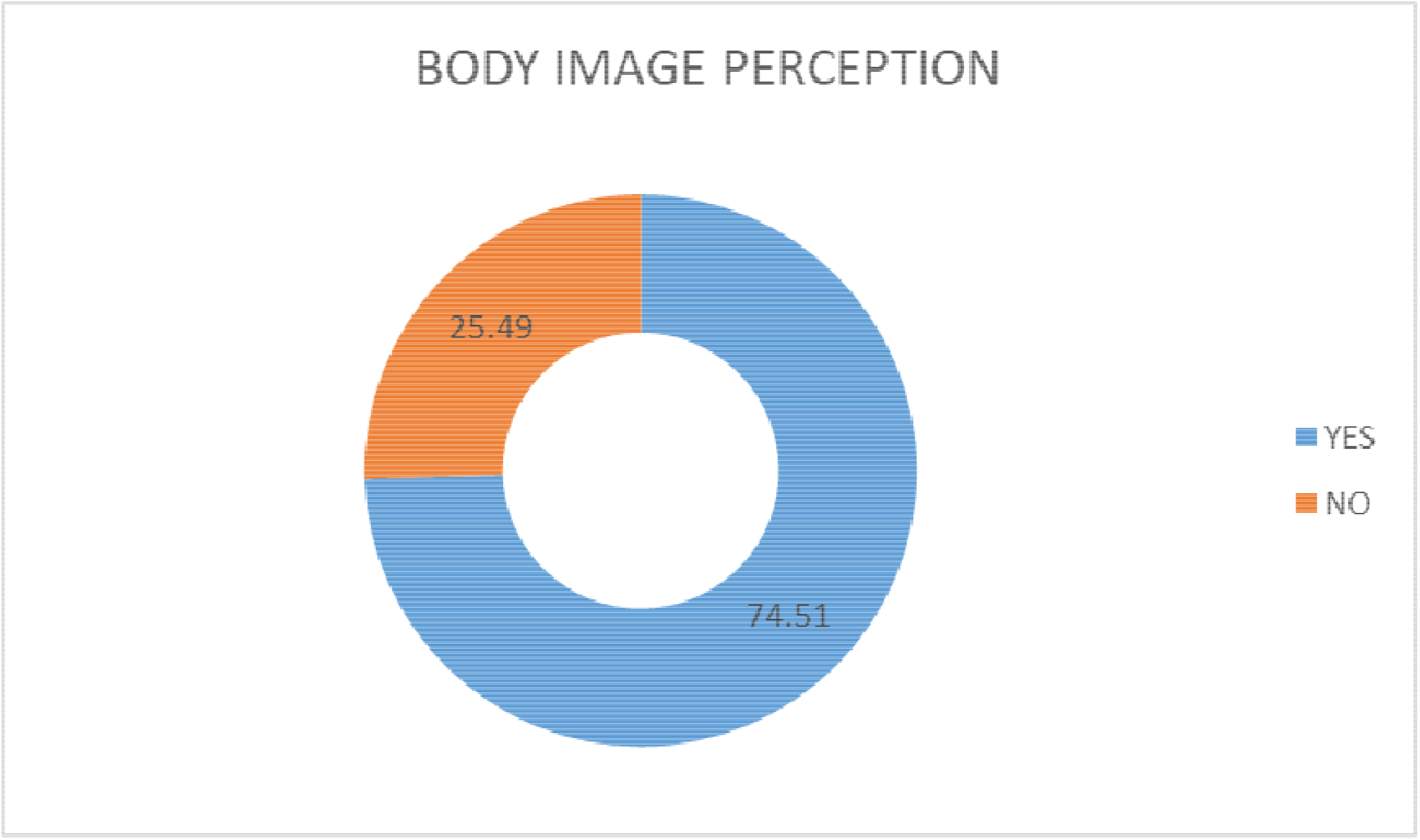

## Discussion

It is evident here that mere exercising has increased total score of participants tremendously and it has helped them overcome their depression and become happy, filled with self-esteem and being satisfied with themselves and life. The statistically significant change seen in table 3 in overall total score of 62.53% is a quite enough proof for that.

Relation with Age as seen in Table 4. The improvement in total score was found to be inversely proportionate with the age. Having said this the participants with younger age group (17-30) showed best improvement of 72.46 % which is statistically significant which results are similar to a review done by Pascoe et al. (23) and another systemic review by Andermo et al. (24) but one cohort study by Bell et al. (25) could only find protective association and not strong evidence. The oldest age group (60-75) showed improvement of only 10.59%, which is not statistically significant. It can be due to the fact that older participants are already having higher self-esteem and satisfaction due to their age and other life experiences compared to (26) than the younger participants (27). As the data suggests, Baseline score of elder participants (11) were already higher than the current-higher score of younger participants.

Table % depicts that Males showed better improvement in total score about 76.70% as also seen in Currier et al. (28), more than double the gain observed in females of 35.83% similar to Manaf et al. (2). In this research which are similar results to Craft et al. (29) and Ohrnberger et al. (5) both changes are statistically significant. It can be due to presence of higher male hormone Testosterone (30) resulting in high Muscle growth and louder manifestations of effects of this hormones on the body. But it was seen that baseline score was slightly higher in females than in males.

When it come to the level of education as seen in Table 6 the improvement of total score was as in age was observed to be inversely proportional to the level of education the participants had received. The changes were found to be statistically significant in all different education levels. Participants who only studied till Secondary education showed an increase in total score by 191.8% whereas participants who Post-graduated showed only difference of only 42.64%. But an important point here can be seen in the baseline score. The Baseline score of Post-graduated participants was around 80% higher than the baseline score of participants with Secondary education. It means education increases happiness and shows a better mental health as shown by different articles (31) and papers as Mowisch et al. (32). But a study published in British journal of psychiatry (33) disagrees. This evidently suggests that Education increases the quality of life by multiple folds and that every child should get education so that it can make its own future by its own hands and live life with the quality that it deserves. Another reason behind this could be lack of long term vision towards future, focus on the present and being happy in small things among Secondary education group which may have increased the total score of present time.

Target achievers vs just exercising (non-target achievers/others) in Table 7 show that the the target achiever group obviously showed better and statistically significant improvement in their total score (22.84%) than the other group (12.28%) which is also statistically significant, similarly to some articles (15) (34) This can be due to the reason that after achieving their target, participants fill accomplished and that boosts their self-esteem by multiple folds and with that self-esteem increases; ultimately increasing their happiness. All of the reasons stated above are proof that the improvement in Target achieving group was better than the just exercising.

Maximum participants (51.96%) agreed to the fact that their parents influenced them most in their life and controversially it was seen here that only (2.94%) participants said that Acting community influenced them most here. (35), also Social media was an important factor (36)People were also influenced by their other family members and some participants even said that God influenced them the most. Chart 1 is sufficient to denote this.

As it can be seen in Chart 2 Body image as an important factor as Almost 74.51% participants in the study agreed to the fact that they thought body shape as an important factor in their perception of anybody similarly to what is mentioned by some articles e.g. (37), (38) and some papers like Harris et al. (39) 25.49% participants said that they thought that personality of anyone and how they would be socially, is completely unrelated with their body shape and that, they were no related at all. What we need to do in the society is to increase this number from 25.49% to more than 90% preferably 100%, to increase the acceptance of all types of people out there and to decrease FAT-SHAMING / THIN-SHAMING and decrease the level of depression and increase the levels of acceptance and self-esteem in people.

### What worked and what didn’t

Data of 102 participants is generally not that big but to derive correlations, it’s a good idea to begin with this sample size. The data is sufficient to conclude that exercising increases not only physical health but mental health also and it causes psychological changes like increased self-esteem, increased satisfaction and increased happiness, thus increasing total life quality. It is evident here that in every demographic group whether female or male, whether young or old, whether well-literate or ill-literate, all participants show increase in their psychological state and even if they don’t show improvement then also this state do not decline or deteriorate. The other important thing here to point is that the study was a Cross-sectional study and not a cohort study. That being said, the participants were told to answer the questionnaire two times in which they had to answer once recalling the time they didn’t use to do exercise. This might have led to **“Recall Bias”**. Other thing important to note is that the Hamilton & Beck’s Depression Inventory is a standard questionnaire for depression meaning that it is used in patients with depression supposedly. Here some questions from the inventory become irrelevant and cannot be answered as the participants are normal people going to gym for exercising.

### Limitations

This study was only done on a **small sample** in an **Urban area** as a **Cross-sectional study** with **Non-Random Sampling**. This study did not consider other forms of physical activity apart from regular exercise in the gym. The research did not include participants from rural areas who might have a different profile.

## Conclusion

This results show that exercising certainly improves self-esteem, satisfaction and helps to reduce depression. This also suggest that we need to increase awareness in people about body image and acceptance. This shows that there is more improvement when the person has achieved their target. And it also suggests that people who do exercise regularly achieve their target definitively and this gives them a feeling of accomplishment thus helping them being more positive and helping others to be the same. People in higher age groups showed to have a better baseline score and therefore showed less improvement, which suggests that the elder people without any socio-economical condition live a relatively stress free life. This also suggests the importance of life experiences in daily life and “being happy at the end of the day”. People who obtained higher education levels demonstrated a better baseline score and therefore showed relatively less improvement, suggestive of importance of exercising, education on self-esteem and self-satisfaction. Males have shown better improvement (more than double) than Females. This important aspect of the study can be due to hormonal and body physiological difference between males and females. This shows that despite being human, each and every body reacts with different elements quite differently.

## Data Availability

All data produced in the present study are available upon reasonable request to the authors

## Acknowledgement

- The authors would like to mention that the project was not funded and there is no conflict of interest
- The authors would like to mention and are thankful to the gym where the study was conducted.

## Recommendations

It can be recommended with this results that Physical therapy and exercise can be included in standard therapeutic protocols of various mental health problems like Depression. The concerned agencies could carry out similar studies with higher sample size to derive better evidences and better policy making. The researchers recommend a prospective “cohort study” with random sampling from Urban and Rural areas with a large sample size including those who are engaged in other forms of activities as well as a control group. Society should be made aware with Body-image and that the effects of that on self-esteem, satisfaction and depression and then awareness against body-shaming should be brought.

## Notes

### Competing Interest Statement

The authors have declared no competing interest.

### Funding Statement

This study did not receive any funding

### Author Declarations

IRB of SMT. NHLMMC (Shrimati Nathiba Hargovindas Lakhamichand Municipal Medical College) gave Ethical approval for this work

